# Controlling biomedical devices using pneumatic logic

**DOI:** 10.1101/2024.01.24.24301744

**Authors:** Shane Hoang, Mabel Shehada, Konstantinos Karydis, Philip Brisk, William H. Grover

## Abstract

Many biomedical devices are powered and controlled by electrical components. These electronics add to the cost of a device (possibly making the device too expensive for use in resource-limited or point-of-care settings) and can also render the device unsuitable for use in some environments (for example, high-humidity areas like incubators where condensation could cause electrical short circuits, ovens where electronic components may overheat, or explosive or flammable environments where electric sparks could cause serious accidents). In this work, we show that pneumatic logic can be used to power and control biomedical devices without the need for electricity or electric components. Originally developed for controlling microfluidic “lab-on-a-chip” devices, these circuits use microfluidic valves like transistors in air-powered logic “circuits.” We show that a modification to the basic valve design—adding additional air channels in parallel through the valve—creates a “high-flow” valve that is suitable for controlling a broad range of bioinstruments, not just microfluidics. As a proof-of-concept, we developed a high-flow pneumatic oscillator that uses five high-flow Boolean NOT gates arranged in a loop. Powered by a single constant vacuum source, the oscillator provides five out-of-phase pneumatic outputs that switch between vacuum and atmospheric pressure every 1.3 seconds. Additionally, a user can adjust the frequency of the oscillator by squeezing a bellows attached to one of the pneumatic outputs. We then used the pneumatic oscillator to power a low-cost 3D-printed laboratory rocker/shaker commonly used to keep blood products, cell cultures, and other heterogenous samples in suspension. Our air-powered rocker costs around $5 USD to build and performs as well as conventional electronic rockers that cost $1000 USD or more. This is the first of many biomedical devices that can be made cheaper and safer using pneumatic logic.

## Introduction

From a lowly lab shaker to a lifesaving ventilator, biomedical devices are ubiquitous throughout the biosciences and medicine. But the widespread use of these valuable tools is often slowed by the cost of these devices. Much of this cost can be attributed to the electronic control hardware (computers, microcontrollers, power supplies, actuators, and so on) that operates the devices. Eliminating this electronic control hardware could make important biomedical devices more feasible for use in resource-limited or point-of-care settings.

In this work, we show that biomedical devices can be made dramatically less expensive by using air (not electricity) to control them. We accomplished this using a pneumatic logic “circuit” that uses air-powered microfluidic valves to serve the role that transistors play in electronic logic circuits. Originally developed for controlling liquids in microfluidic chips [1], these monolithic membrane valves have been used in a variety of pneumatic logic circuits for controlling liquid flow in microfluidic “lab-on-a-chip” devices [2–11]. However, the volumes of air controlled in these circuits are usually on the microliter scale—far too small for controlling most biomedical devices. To solve this problem, we developed “high flow” monolithic membrane valves that use multiple parallel channels to control much larger volumes of air than conventional valves [12].

For this initial demonstration of pneumatic-logic-based biomedical device control, we chose to target the wide variety of biomedical devices that utilize periodic or oscillatory motions. For example, intermittent pneumatic compression (IPC) devices periodically squeeze a patient ‘s legs to encourage blood flow and counteract the formation of clots [13–15]; laboratory rockers and shakers use repetitive tilting or swaying motions to keep blood and cell cultures in suspension [16]; and ventilators move air into and out of the lungs [17, 18]. Devices like these typically use electricity, motors or pumps, and computers or microcontrollers to create and control these periodic motions. All of this hardware adds considerable expense and complexity to these devices. For example, while the IPC stockings worn by patients are inexpensive enough to be single-use and disposable, the electromechanical hardware used to send periodic pneumatic signals to the stockings cost thousands of dollars per unit [19]; this complicates the widespread use of IPCs in care facilities and homes. Likewise, blood banks can need large numbers of lab rockers to keep blood products suspended and oxygenated and avoid coagulation; purchasing, powering, and maintaining all this electromechanical equipment can be a significant burden for health facilities in resource-limited settings. Additionally, electronic lab rockers and shakers may be unsuitable for use in some environments, such as high-humidity incubators (where moisture might encourage electrical short-circuits or corrosion), refrigerators and freezers (where condensation can damage electrical circuits), ovens (where overheating can damage motors and microprocessors), and flammable or oxygen-rich environments (where an electrical spark could cause a fire or explosion).

To demonstrate that pneumatic logic circuits can provide periodic or oscillatory signals for controlling biomedical devices, we developed a high-flow version of a pneumatic logic oscillator originally created for controlling microfluidic chips [20]. Our oscillator is powered by a single constant vacuum source and provides five pneumatic outputs that automatically and continuously alternate between vacuum and atmospheric pressure; these outputs can in turn be used to power and control biomedical devices. To demonstrate this, we used our high-flow pneumatic oscillator to control a 3D-printed laboratory rocker/shaker device. Our air-powered rocker costs about $5 USD to make and performs as well as conventional electronic rockers that cost $1000 USD or more; it is the first of many biomedical devices that can be made cheaper and safer using pneumatic logic.

## Materials and methods

### Pneumatic oscillator design and operation

Our pneumatic logic circuits use monolithic membrane valves to create logic gates. As shown in Figure 1A, these valves consist of a featureless commercially-produced polydimethylsiloxane (PDMS) silicone rubber sheet sandwiched between two rigid layers containing etched or engraved channels. In this work, we used engraved acrylic plastic sheets for the channel layers. A valve is formed wherever an engraved chamber in one acrylic sheet is located directly across the PDMS sheet from a gap in a channel in the other acrylic sheet. Using multiple channels in parallel creates a “high-flow” valve suitable for containing larger air flows than are typically encountered in microfluidics [12].

**Figure 1:**
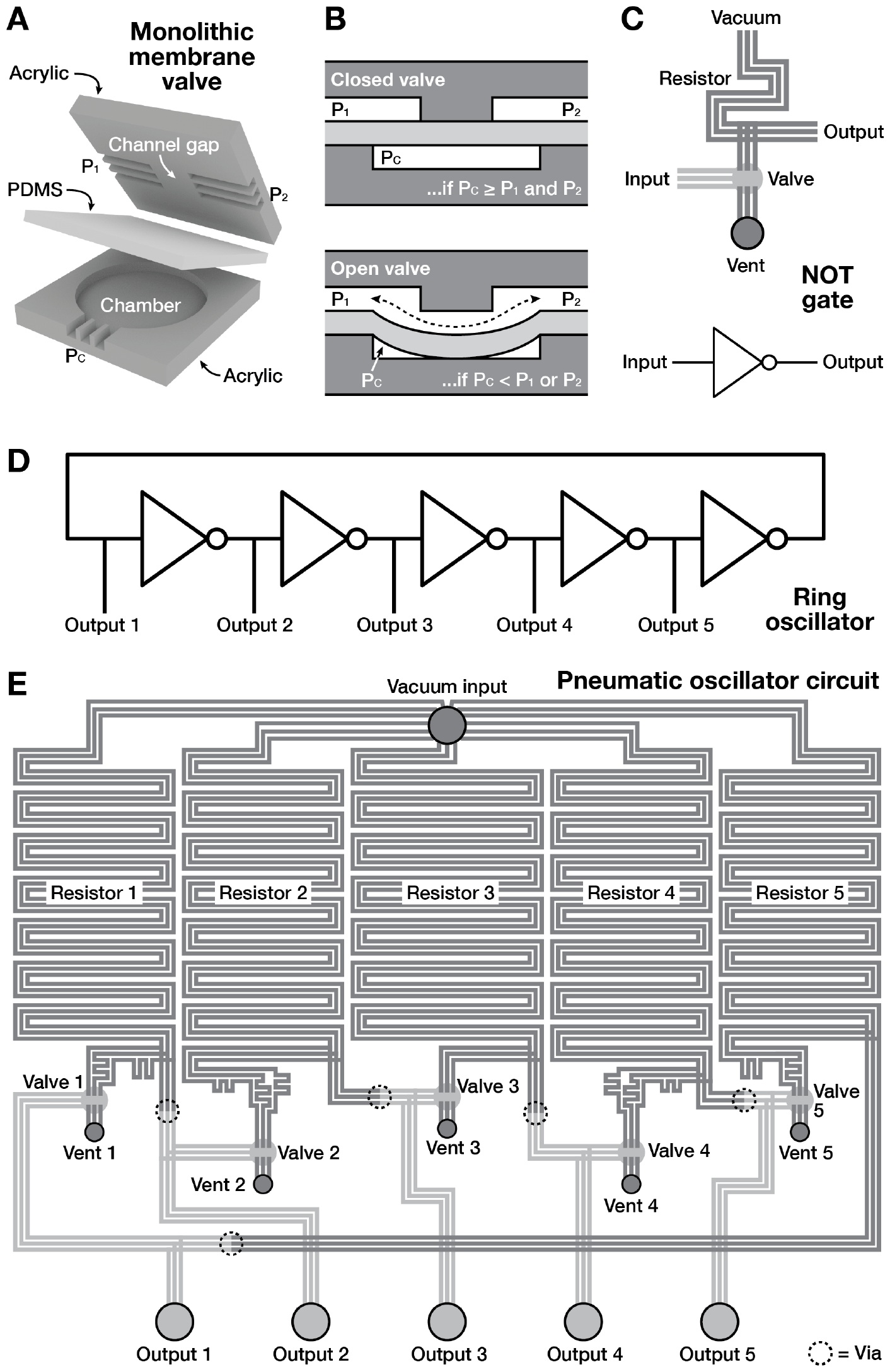
Exploded (A) and cross-section (B) views of a single high-flow monolithic membrane valve. Combining a valve with a vent hole and a long pneumatic resistor channel creates a Boolean NOT gate (C) represented by the symbol shown. An odd number of NOT gates arranged in a loop creates a ring oscillator (D). Design of a pneumatic ring oscillator with five outputs (E). When a constant vacuum is applied to the vacuum input, the five outputs automatically and continuously oscillate between vacuum and atmospheric pressure, all at the same frequency but offset in time (out of phase).

**Figure 2:**
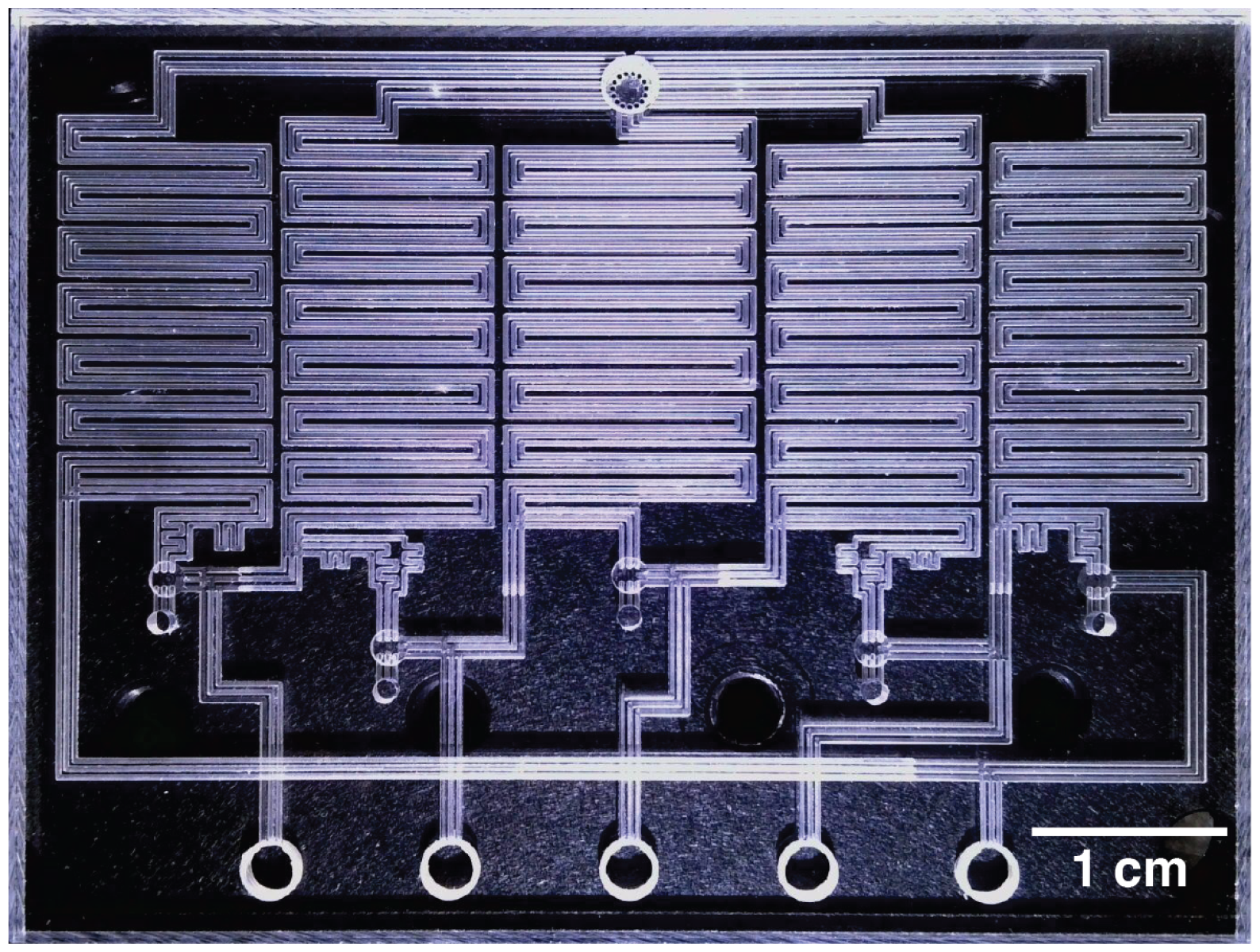
Photograph of a completed high-flow pneumatic oscillator. A single constant vacuum source at the top input powers five outputs at the bottom that automatically oscillate between vacuum and atmospheric pressure.

The cross-sectional view through a single valve in Figure 1B shows that these valves are normally closed: when the same pressure is applied to both the valved channels and the chamber, the PDMS sheet seals against the gap in the valved channels and no air flows through the valve. However, when a vacuum is applied to the chamber, the PDMS sheet stretches into the chamber and creates a path for air to flow across the gap in the valved channels (the dotted arrow in Figure 1B), and the valve opens. More generally, for a valve with pressures *P*_1_ and *P*_2_ at the two ends of the valved channel and pressure *P*_*C*_ at the chamber:

- If *P*_*C*_ *≥ P*_1_ and *P*_*C*_ *≥ P*_2_, then the valve will be closed (Figure 1B, top)
- If *P*_*C*_ *< P*_1_ or *P*_*C*_ *< P*_2_, then the valve will be open (Figure 1B, bottom); air will flow from channel 1 to channel 2 as long as *P*_1_ *> P*_2_, or from 2 to 1 as long as *P*_2_ *> P*_1_

Finally, by arbitrarily assigning a logical meaning of TRUE for a vacuum and FALSE for atmospheric pressure, binary information can be encoded and manipulated as different air pressure levels inside the device. In this manner, pneumatic logic circuits can be constructed by connecting valves together using air channels.

Our pneumatic oscillator circuit uses Boolean NOT gates; these fundamental logic gates output the opposite of their input (so if the input is TRUE then the output is FALSE, and if the input is FALSE then the output is TRUE). Figure 1C shows the design of the valve-based pneumatic NOT gate used in this study; this design is based on one originally developed for microfluidic chip control [3] but is implemented here using our high-flow valves. The NOT gate is powered by a constant vacuum source that pulls air through a long section of channel that serves as a pneumatic resistor. The output of the resistor splits; one end is connected to a valved channel, and other end is connected to the output of the gate. The other end of the valved channel is connected to a vent (a drilled hole that connects the contents of the channel to the atmosphere). Finally, the valve chamber is connected to the input of the gate.

When atmospheric pressure (FALSE) is applied to the input of the pneumatic NOT gate in Figure 1C, the valve chamber remains at atmospheric pressure and the valve remains closed. This means that air from the output can flow through the resistor to the vacuum source; this creates a vacuum (TRUE) at the output, as expected according the definition of the NOT gate. Conversely, when vacuum (TRUE) is applied to the input of the NOT gate, the valve chamber is under vacuum and the valve opens. This creates a low-resistance path from the output through the valve to the vent, effectively placing the output at atmospheric pressure. While the vacuum source still pulls some air from the output, the different resistances of the two flow paths (the low-resistance path to the atmospheric vent versus the high-resistance path to the vacuum source) ensure that the output is at atmospheric pressure (TRUE), again as expected for a NOT gate. In this manner, the pneumatic NOT gate aways outputs the opposite of its input.

When an odd number of NOT gates are connected in a loop as shown in Figure 1D, the resulting circuit is a ring oscillator. This unstable circuit automatically and continuously alternates the outputs between TRUE and FALSE. To understand why, imagine that the circuit starts with output 1 = TRUE. This is negated by the first NOT gate, so output 2 = FALSE, which makes output 3 = TRUE, which makes output 4 = FALSE, which makes output 5 = TRUE. This is negated by the final NOT gate to FALSE, which is then connected to output 1. This effectively flips output 1 from its original TRUE to FALSE, and it also causes all subsequent outputs to flip as well, then the cycle repeats. In this way, the values of all five outputs automatically and constantly flip between TRUE and FALSE, with output-flipping propagating like a wave traveling continuously around the loop.

By using five pneumatic NOT gates to build a ring oscillator, we created the pneumatic oscillator shown in Figure 1E. Based on a design developed by Duncan *et al*. for controlling small volumes of air to operate microfluidic devices [20], our version of the oscillator uses high-flow valves to control larger volumes of air. A single vacuum input at the top of the oscillator design powers all five NOT gates. Vias (holes punched through the PDMS membrane; dotted circles in Figure 1E) allow pneumatic signals to pass from one side of the membrane to the other. A photograph of a fabricated pneumatic oscillator is shown in

### Pneumatic oscillator fabrication

The pneumatic oscillator was designed in Adobe Illustrator (file available as online *Supplementary Information*) and fabricated using a desktop CNC mill (Bantam Tools; Peekskill, New York) to engrave all device features into two acrylic plastic sheets (6.35 cm wide, 5.08 cm long, and 3 mm thick). All channels were engraved to a width and depth of 450 *μ*m, and valve chambers were milled out to a circular shape with a diameter of 3 mm and a depth of 450 *μ*m. Vents (2 mm diameter) and outlets (4 mm diameter) were milled through the entire thickness of the acrylic sheet. The single vacuum input was engraved as a circle with a diameter of 4 mm and depth of 2.3 mm; this leaves a 0.25 mm region of acrylic between the bottom of the vacuum input and the other side of the acrylic (as 0.45 mm was already milled out on the other side for the vacuum channels). Small holes (0.45 mm in diameter) were then milled through this 0.25 mm region; this provides a path for air to flow from the vacuum channels to the vacuum input while also preventing the polydimethylsiloxane (PDMS) membrane from being drawn into the opening and inadvertently blocking the air flow after the device is bonded. Finally, the inlet and outlet holes were tapped with 10-32 threads.

To bond the pneumatic oscillator, the two acrylic sheets were first rinsed with 99.5% isopropyl alcohol, then rinsed with purified water, then submerged for 20 minutes in a 5% (v/v) solution of 3-aminopropyltriethoxysilane (Sigma-Aldrich, St. Louis, MO) diluted in purified water. Next, a 250 *μ*m thick sheet of commercially-produced polydimethylsiloxane (PDMS) silicone rubber (HT-6240; Rodgers Corporation/Bisco Silicones, Carol Stream, IL) was cut to the dimensions of the acrylic sheets. A 3 mm diameter biopsy punch (Electron Microscopy Sciences, Hatfield, PA) was used to punch holes in the PDMS membrane to form vias. The bonding surfaces of the PDMS sheet and the acrylic sheets were then treated for one minute using a corona treater (BD-20AC; Electro-Technic Products, Chicago, IL), after which all the layers were assembled together into the acrylic-PDMS-acrylic sandwich shown in Figure 1. The oscillator was then clamped overnight to give the PDMS-acrylic bonds time to strengthen. Finally, threaded barbed tubing connectors were screwed into the input and output connectors.

### Pneumatic oscillator testing

To characterize the performance of the high-flow pneumatic oscillator, we used tubing to connect its vacuum input to the laboratory building vacuum supply ( *−* 60 kPa) and connect the five output connections to five small plastic bellows intended for use as fluid dispensers (“Yueton” droppers/pipettes; amazon.com). Each output connection was also connected to a custom-built open-source multichannel pressure logger to record the pressure at each output during device operation. The logger uses an Arduino Nano microcontroller and a custom printed circuit board (PCB) to acquire data from up to eight digital pressure sensors (MPX4250DP; NXP Semiconductors, Austin, TX) and relay these pressure measurements via USB to a computer running a custom Python data acquisition program. Printed circuit board design files and Arduino and Python code for the pressure logger are available as online *Supplementary Information* and in the pressure logger ‘s GitHub repository [21].

To demonstrate using our pneumatic oscillator to operate a typical biomedical device, we designed and fabricated the 3D-printed laboratory rocker/shaker shown in Figure 3. We chose a rocker because of their importance in many different research and medical settings. For example, in blood banks, rockers are needed to constantly agitate platelets from donation until transfusion [22]. Many agglutination assays that are used to detect antibodies and diagnose diseases also rely on rockers. Rockers and shakers are also used in solid-liquid and liquid-liquid extractions, sample emulsification, staining and destaining samples like electrophoretic gels and blots, and preventing sedimentation in a wide range of heterogeneous samples. Conventional electronic rockers typically cost $1000 USD or more, which limits their widespread use, especially in resource-limited settings. Consequently, a low-cost rocker powered by our pneumatic oscillator could be a valuable tool for researchers and clinicians around the world. Additionally, a fully pneumatic (non-electronic) rocker could be used safely around flammables, explosives, high humidity, and other conditions that would be incompatible with conventional rockers powered by electricity.

**Figure 3:**
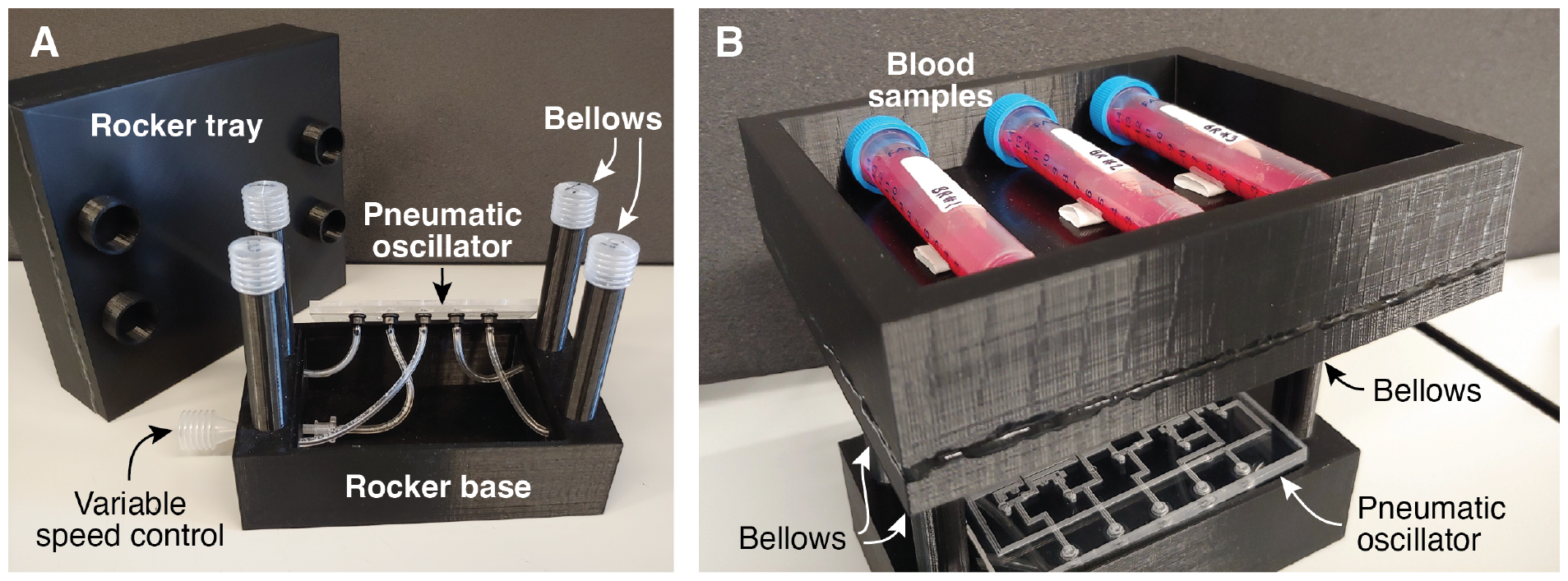
(A) This 3D-printed laboratory rocker/shaker is powered by our pneumatic oscillator. Four plastic bellows are connected to the pneumatic oscillator ‘s outputs by tubing. (B) A tray sits on top of the bellows and holds samples for agitation.

Our 3D-printed rocker contains four small plastic bellows (visible in Figure 3A) that are connected via tubing to four of the outputs on our pneumatic oscillator (outputs 1, 2, 4, and 5). The four bellows are mounted so that the rocker ‘s moving tray rests on top of the bellows, as shown in Figure 3B. A hollow base provides room for the pneumatic oscillator. The fifth output of the pneumatic oscillator is connected to an additional bellows that serves as a variable speed control. The rocker was designed using Solidworks and fabricated using a low-cost 3D printer (Ender-3; Creality, Shenzhen, China) using PLA filament.

To test the performance of the pneumatic-oscillator-powered rocker, three 15 mL Falcon-style centrifuge tubes were each loaded with 5 mL of fresh whole bovine blood (Na-citrate anticoagulant; Lampire Biological Laboratories, Pipersville, PA) before placing the tubes side-by-side on the blood rocker as shown in Figure 3B. The pneumatic oscillator ‘s vacuum input was then connected to the laboratory building vacuum supply, and the blood rocker was operated nonstop for seven days. Once per day the tubes were gently removed and photographed to check for signs of separation in the blood. During this 7-day period, a fourth Falcon tube with 5 mL of whole bovine blood was left upright at room temperature to act as a control for comparison.

## Results

Figure 4 shows typical results from operating the pneumatic oscillator nonstop for over two days. A full cycle of the oscillator is visible when viewing 1.5 seconds of data (Figure 4A): starting arbitrarily on the left with TRUE for output 1, this signal is inverted to FALSE for output 2, which is inverted to TRUE for output 3, then FALSE for Output 4, then TRUE for Output 5, then FALSE for output 1, then TRUE for output 2, then FALSE for output 3, then TRUE for output 4, and finally FALSE for output 5, then the cycle repeats. Each full cycle takes about 1.2 seconds.

**Figure 4:**
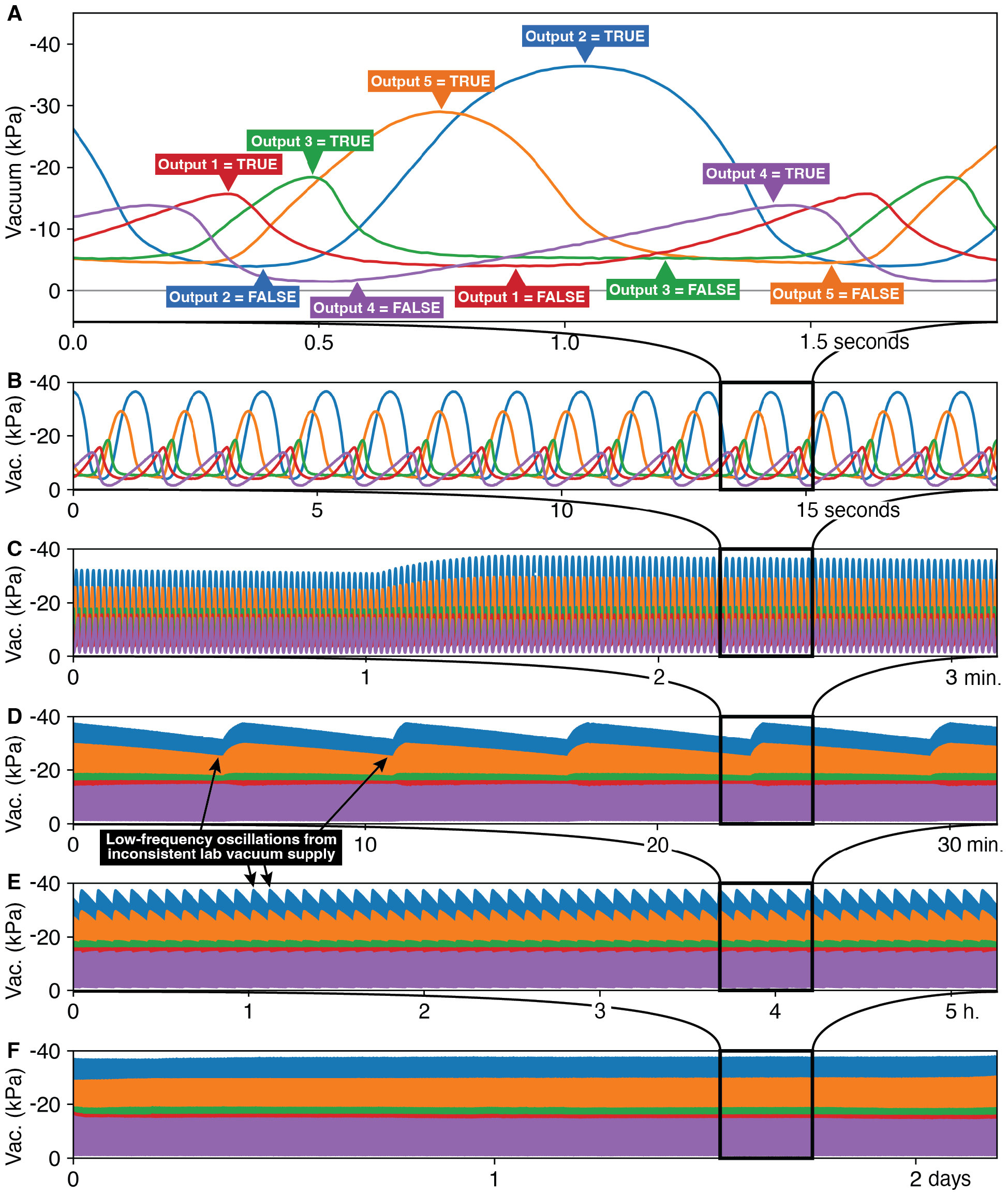
Vacuum pressure measured at each of the five pneumatic oscillator outputs versus time during two days of nonstop operation, zooming out by successive factors of ten to view 1.5 seconds (A), 15 seconds (B), 3 minutes (C), 30 minutes (D), 5 hours (E), and the full 2 days (F). Pressures are relative to atmospheric pressure (0 kPa) and plotted so that higher-magnitude vacuums are higher on the Y-axis.

Figure 4A also reveals that the five outputs of the pneumatic oscillator reach different maximum vacuums during the oscillation cycle. Output 2 reaches the the highest vacuum at *−* 35 kPa, followed by output 5 at *−* 30 kPa, and the remaining three outputs reach maximum vacuums of between *−* 15 and *−* 20 kPa. These variations between output pressures could be caused by differences in the channel lengths between the different NOT gates; for example, the channel that connects the output of NOT 5 to the input for NOT 1 is considerably longer than the channels that connect the other NOT gates, and this additional resistance could slow the flow of air between NOT gates 5 and 1 and possibly decrease the maximum vacuum reached on output 1. Additionally, small gate-to-gate variations in valve behavior could manifest themselves as different output pressures from the different NOT gates. Regardless of the cause of the differences in output vacuums, zooming out by a factor of ten (Figure 4B) shows that the maximum output vacuums reached by the five oscillator outputs remain consistent.

Zooming out further (Figure 4C, D, and E) reveals low-frequency oscillations in the magnitudes of the output pressures, with pressures abruptly rising and then slowly falling every six minutes. We found that these low-frequency oscillations were caused by regular variations in the pressure of the laboratory building ‘s central “house vacuum” supply used to power the oscillator. These oscillations can be easily eliminated by using a more consistent vacuum supply; however, since inconsistent vacuum supplies may be unavoidable in many settings, we continued to use the laboratory building vacuum supply to better understand the effect of inconsistent vacuum supplies on oscillator operation. Finally, zooming out by one more factor of ten (Figure 4F) shows that (apart from the low-frequency variation caused by the inconsistent laboratory vacuum supply) the maximum vacuum at each of the pneumatic oscillator ‘s five outputs stays consistent over the entire two-day-long experiment.

We also analyzed the frequency stability of the pneumatic oscillator over this two-day run. During the first three hours, the average oscillation frequency was 0.749 Hz or an average period of 1.335 s per cycle. The last three hours had an average oscillation frequency of 0.779 Hz or 1.283 s/cycle. This means that over two days of constant operation (during which the oscillator completed around 140,000 cycles and 700,000 valve openings and closings) the oscillation frequency of the pneumatic oscillator changed by only 3.9%. While the frequency stability of our high-flow pneumatic oscillator is far lower than the stability of electronic oscillators for *e*.*g*. timekeeping applications, a few percentage points of frequency variation over days of operation is acceptable for many biomedical device applications.

To test our pneumatic-oscillator-controlled laboratory rocker/shaker shown in Figure 3, we placed samples of whole bovine blood on the tray and turned on the vacuum supply to the oscillator. The pneumatic oscillator began rocking the blood samples in a gentle back-and-forth motion. We left the blood samples on the rocker for seven days, gently removing them once per day to photograph their contents. A “control” tube of blood was left stationary for seven days and photographed daily. Figure 5 shows that while the stationary “control” blood was visibly separating after one day and fully separated into cell and plasma layers after seven days, the three blood samples on our pneumatic oscillator rocker showed no visible changes and remained in suspension throughout the entire seven-day experiment.

**Figure 5:**
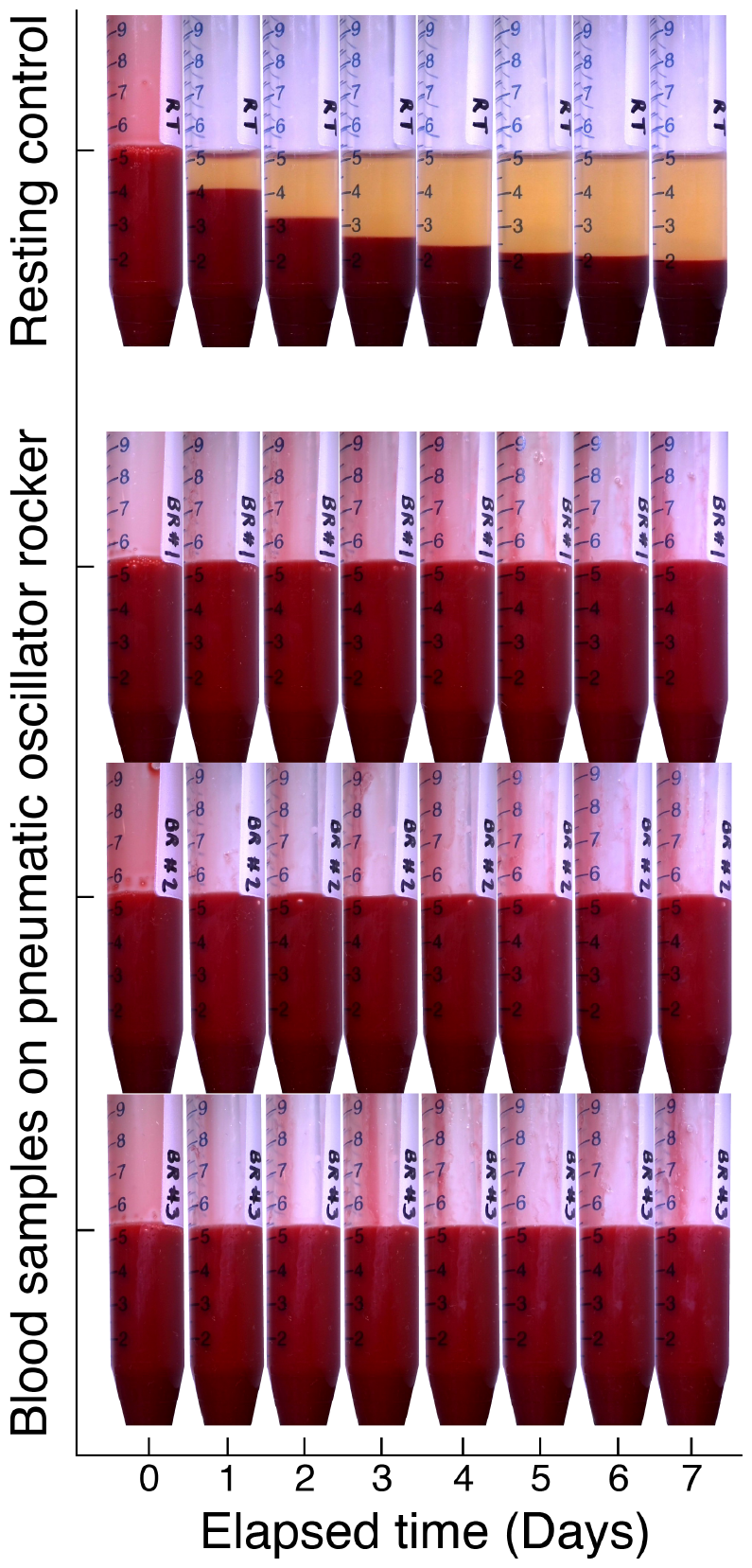
Photographs of bovine blood samples over seven days spent either sitting at rest (top sample) or on our 3D-printed air-powered pneumatic oscillator rocker (bottom three samples). While the resting control sample quickly separated into cell and plasma layers, the rocker successfully kept its three samples in suspension for the entire seven-day experiment.

While testing the pneumatic oscillator, we observed that manually holding one of the five output bellows in a compressed state (as shown in the top-right of Figure 6) noticeably increases the oscillation speed of the pneumatic oscillator. We attribute this to the change in the volume of air that the pneumatic oscillator is depressurizing and pressurizing with each cycle. A manually-compressed bellows contains less air than an expanded one, so when the pneumatic oscillator applies vacuum to the bellows, it takes less time to decompress the smaller volume inside the compressed bellows than it takes to decompress the larger volume inside the expanded bellows. Similarly, the manually-compressed bellows re-pressurizes faster than a freely-moving bellows when the oscillator applies atmospheric pressure to the bellows. Or, to use an electrical analogy, a free-moving bellows is like a large capacitor that takes more time to charge and discharge in an electric circuit, and a manually-compressed bellows is like a small capacitor that takes less time to charge and discharge.

**Figure 6:**
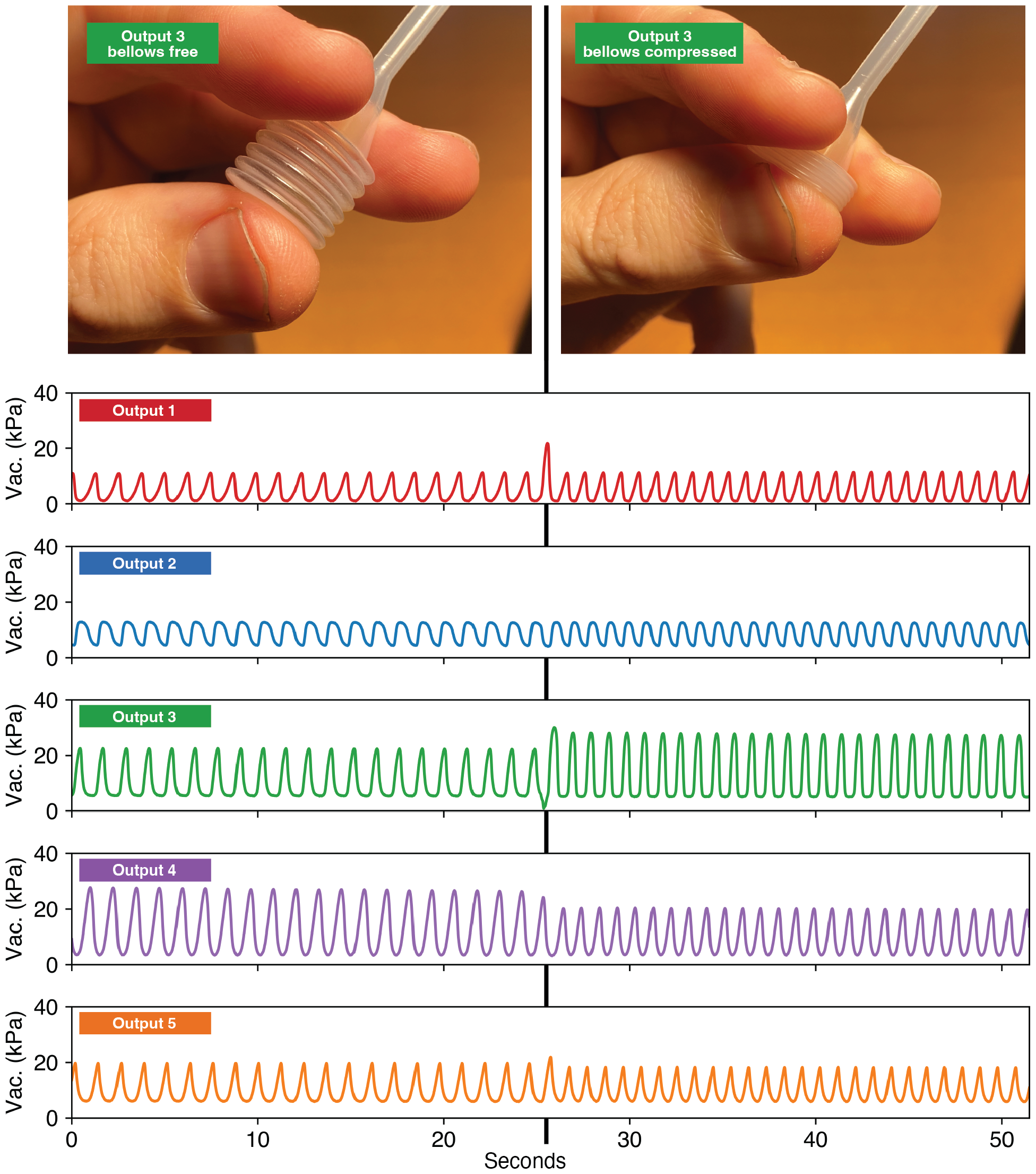
Vacuum pressure measured inside each of the five outputs of the pneumatic oscillator, with the “variable speed control” bellows on output 3 free (0 to 26 seconds) and manually compressed (26 to 50 seconds). Holding the “variable speed control” bellows increases the pneumatic oscillator ‘s speed by 33%.

We then realized that manually-compressed bellows could be used as a sort of “variable speed control” for the pneumatic oscillator. To test this idea, we connected our multichannel pressure logger to our rocker and monitored the pressures in each of the five pneumatic outputs both before and after manually compressing the bellows on Output 3 (the bellows labeled “variable speed control” in Figure 3). The results are shown in Figure 6. During the first 26 seconds, the “variable speed control” bellows is free to expand and contract as usual, and the pneumatic oscillator has a measured frequency 0.798 Hz (or a period of 1.253 s per cycle). Then, we manually held the “variable speed control” bellows in the compressed state for the remainder of the run, and the pneumatic oscillator ‘s frequency immediately increased to 1.048 Hz (a period of 0.954 s per cycle). Manually compressing the “variable speed control” bellows increased the rocker ‘s oscillation speed by 33%. This suggests that the speed of our air-powered rocker can be tailored for a given application simply by adjusting the volumes of the bellows.

## Discussion

Pneumatic logic is not a new idea—pneumatic systems were used to individually control each room ‘s temperature in large office buildings in the late 1800s [23], and player pianos used air to read notes from punched-paper songs in the early 1900s [24]. These systems fell out of favor when transistors and microprocessors made electronic control ubiquitous. However, there remain many applications where avoiding the cost and complexity of electronic hardware can be advantageous. By using monolithic membrane valves like air-powered transistors in pneumatic logic circuits, we have shown that we can control sophisticated biomedical devices without the need for electricity or electromechanical hardware.

The cost savings from pneumatic logic can be significant. We estimate that our pneumaticoscillator-powered rocker contains about $5 USD worth of materials—less than 1% of the cost of a conventional electronic rocker, and also considerably cheaper than existing 3D-printed rockers that do not utilize pneumatic logic [25]. Our rocker does require a vacuum source to power it, but many labs and clinics already have central “house vacuum” available (and our results in Figure 4 show that the pneumatic oscillator continues to function even when the house vacuum supply is unreliable). And in resource-limited settings without central vacuum, a single low-cost air pump (like the sub-$10-USD models used to aerate an aquarium) could easily provide enough air flow to power several pneumatic oscillator rockers.

In addition to cost savings, pneumatically controlled biomedical devices can have safety advantages as well. With no danger of sparks or fires, no damage from moisture or humidity, and no way to generate (or receive) electromagnetic interference, pneumatic-logic-powered devices can be safer than conventional electronic tools in hospital beds, incubators, refrigerators and freezers, medical imagers, operating rooms, and many other settings.

Finally, oscillators and rockers are just the “tip of the iceberg” of biomedical applications for pneumatic logic. Far more complex pneumatic logic circuits are possible—an entire air-powered computer was recently developed for controlling microfluidic “lab-on-a-chip” devices using monolithic membrane valves [11]—so even complex biological and medical devices could be controlled by pneumatic logic.

## Supporting information

Design of pneumatic oscillator in Adobe Illustrator format

Printed circuit board design files and Arduino and Python code for the custom multichannel pressure logger

## Data Availability

All data produced in the present work are contained in the manuscript.

## Acknowledgments

This work was supported by the National Science Foundation ‘s Division of Civil, Mechanical and Manufacturing Innovation (CMMI) under award numbers 1740052 (Philip Brisk), 2046270 (Konstantinos Karydis), and 2133084 (Konstantinos Karydis), the National Science Foundation ‘s Division of Computing and Communication Frontiers (CCF) under award number 2019362 (William H. Grover), and the National Science Foundation ‘s Division of Biological Infrastructure (DBI) under award number 2131428 (William H. Grover).

## Supplementary Information

Online Supplementary Information is also available:

- Design of pneumatic oscillator in Adobe Illustrator format
- Printed circuit board design files and Arduino and Python code for the custom multichannel pressure logger

