## Supplementary material for "Controlling biomedical devices using pneumatic logic": Printed circuit board design files and Arduino and Python code for the custom multichannel pressure logger: pressurama-arduino.ino

```
void setup() {
  Serial.begin(2000000);
}

int averages = 10;

void loop() {
  float sums[] = {0,0,0,0,0,0,0,0};
  for(int measurement = 0; measurement < averages; measurement++) {
    for(int pin = 0; pin < 8; pin++) {
      sums[pin] += analogRead(pin);
    }
  }
  Serial.print("X ");
  for(int pin = 0; pin < 8; pin++) {
    Serial.print(pin);
    Serial.print(":");
    Serial.print(sums[pin] / averages);
    Serial.print(" ");
  }
  Serial.print("Y\n");
}
```
