## Supplementary material for "Controlling biomedical devices using pneumatic logic": Printed circuit board design files and Arduino and Python code for the custom multichannel pressure logger: pressurama-mac.py

```
import serial, os, sys, datetime
outfile = open("out.csv", "a")
port = ""
usb_count = 0
devices = os.listdir("/dev")
for device in devices:
    # if "cu.usbserial" in device:  <-- original
    # Recent versions of MacOS seem to use cu.usbmodem
    # so this version will catch both:
    if "cu.usb" in device:
        port = device
        usb_count += 1
if usb_count == 0:
    sys.exit("No port found")
if usb_count > 1:
    sys.exit("Multiple ports found")
port = "/dev/" + port
data = []
ser = serial.Serial(port, 2000000)
ser.flush()
while True:
    s = ser.readline().decode("utf-8")
    if s.startswith("X") and s.strip().endswith("Y") and \
    s.count("X") == 1 and s.count("Y") == 1 and \
    s.count(" ") == 9 and s.count(":") == 8:
        print(ser.inWaiting(), datetime.datetime.now().isoformat(), end="\t")
        outfile.write(datetime.datetime.now().isoformat())
        tokens = s.split(" ")
        for token in tokens:
            if ":" in token:
                channel = int(token.split(":")[0])
                measurement = float(token.split(":")[1])
                print("%0.1f\t" % (measurement), end="")
                outfile.write("," + str(measurement))
        print()
        outfile.write("\n")
        # outfile.flush()
        # os.fsync(outfile)
```
