## Supplementary material for "Controlling biomedical devices using pneumatic logic": Printed circuit board design files and Arduino and Python code for the custom multichannel pressure logger: readme.txt

Using default Python from python.org

PC version currently has a hard-coded COM port

Dependencies for pressurama data acquirer:
    pip install pyserial

Dependencies for plotters:
    pip install matplotlib
